# Use of magnetic resonance imaging in neuroprognostication after pediatric cardiac arrest: Survey of current practices

**DOI:** 10.1101/2022.01.25.22269764

**Authors:** Juan A. Piantino, Christopher M. Ruzas, Craig A. Press, Subramanian Subramanian, Binod Balakrishnan, Ashok Panigrahy, David Pettersson, John A. Maloney, Arastoo Vossough, Alexis Topjian, Matthew P Kirschen, Lesley Doughty, Melissa G. Chung, David Maloney, Tamara Haller, Anthony Fabio, Ericka L. Fink, the POCCA Investigators

## Abstract

**Background:** Use of MRI as a tool to aid in neuroprognostication after cardiac arrest (CA) has been described, yet details of specific indications, timing, and sequences are unknown. We aim to define the current practices in use of brain MRI in prognostication after pediatric cardiac arrest.

**METHODS:** A survey was distributed to pediatric institutions participating in three international studies. Survey questions related to center demographics, clinical practice patterns of MRI after CA, neuroimaging resources, and details regarding MRI decision support.

**RESULTS:** Response rate was 31% (44/143). Thirty-four percent (15/44) of centers have a clinical pathway informing the use of MRI after CA. Fifty percent (22/44) of respondents reported that an MRI is obtained in nearly all CA patients and 32% (14/44) obtain an MRI in those who did not return to baseline neurological status. Sixty-four percent of centers (28/44) obtain an MRI greater than 72 hours after return of spontaneous circulation. Poor neurologic exam was reported as the most common factor (91% [40/44]) determining the timing of the MRI. Conventional sequences (T1, T2, FLAIR, and diffusion weighted imaging/apparent diffusion coefficient) are widely used. Advanced imaging techniques, such as MR spectroscopy, diffusion tensor imaging and resting state functional MRI are less commonly used.

**CONCLUSIONS:** Conventional brain MRI is a common practice for prognostication after CA. Advanced imaging techniques, are used infrequently. The lack of standardized clinical pathways and variability in reported practices support a need for higher-quality evidence regarding the indications, timing, and acquisition protocols of clinical MRI studies.

## INTRODUCTION

Pediatric cardiac arrest (CA) is a relatively uncommon event yet continues to be a major public health concern. Epidemiological studies have described a prevalence of nontraumatic out-of-hospital CA (OHCA) of 8 per 100,000 person years and a prevalence if in-hospital CA (IHCA) of 0.77 per 1,000 hospital admissions. Mortality rates range from 40-60% for IHCA with greater than 90% for OHCA.^1-5^ In those who survive to hospital discharge, rates of survival with favorable neurologic outcome are 4-16%^6,7^ for OHCA and 80-96%^3, 5^ for IHCA.

Multiple monitoring modalities including electroencephalography (EEG), laboratory tests, and neuroimaging, are used to aid management and prognostication after CA.^8-11^ However, no single modality has sufficient accuracy to facilitate neurological prognostication after return of spontaneous circulation (ROSC).^12^ In recent years, brain magnetic resonance imaging (MRI)-based biomarkers have been increasingly recognized as promising tools to evaluate children after CA. Recent studies have shown that conventional MRI sequences (T1, T2, and diffusion-weighted imaging [DWI]) can accurately identify hypoxic-ischemic injury and can be used to assist in prognostication.^13-19^ Additionally, more advanced MRI techniques, such as MR spectroscopy (MRS), are able to measure the concentration of metabolites that serve as markers of injury and may also have prognostic value.^10,20,21^ Quantitative whole brain white matter fractional anisotropy (FA) measurements utilizing diffusion tensor imaging (DTI) are associated with long term neurological outcome in patients with cardiac arrest.^18^ Resting state functional MRI (rs-fMRI) has also been shown to have prognostic information in cardiac arrest patients who are comatose and have indeterminate prognosis and patients with higher functional connectivity recovered consciousness.^19^

Despite these advances, there is a lack of evidence-based consensus regarding the indications, timing, and MRI sequences most useful in this population, and little is known about current practice. The objective of this study was to assess current practices of post-CA MRI for neuroprognostication at international tertiary care pediatric centers. To achieve our objective, we surveyed pediatric centers to assess practices in four areas: neuro-imaging resources, rationale for MRI, timing of MRI acquisition following CA, and specific MRI sequences used.

## METHODS

### Study design

This was a survey of current practices regarding brain MRI use in children with post-CA. The study was approved by the University of Pittsburgh Institutional Review Board as “Not Human Research” (STUDY20010055). The survey was conducted between May and August 2020. Pediatric hospitals participating in the Personalizing Outcomes after Child Cardiac Arrest (POCCA) study, the Approaches and Decisions after Pediatric Traumatic Brain Injury (ADAPT) trial, and the Prevalence of Acute critical Neurological disease in children: a Global Epidemiological Assessment (PANGEA) study were invited to participate in the survey. All participating sites have expertise in critical care and imaging-related research.^22-24^

### Survey development and distribution

The 16-item survey was developed in consultation with experts in the field of Pediatric Critical Care and Neuroradiology at the University of Pittsburgh and Oregon Health & Science University. After the initial survey was developed, the remainder of the study group provided feedback. The final survey consisted of a multiple-choice questionnaire divided into two sections. The first section contained general information about each institution, the characteristics of each intensive care unit (total number of pediatric intensive care unit [PICU] beds, annual number of pediatric CA), and neuro-imaging resources (number and type of MRI scanners, availability of experts in neuroradiology). The second section focused on MRI practices and presence of an institutional clinical pathway to support clinical decisions surrounding indications of MRI after CA, timing, and MRI sequences used in these patients.

The survey was distributed by the POCCA data coordinating center to all the participating sites. Each site’s medical director or principal investigator was contacted via email and asked to complete the survey, with the assistance of the institution’s neuroradiologist if needed. Follow-up emails were sent every two weeks up to four times. A single survey was completed per institution. If an individual site participated in more than one of the studies used to generate the survey distribution list, the site principal investigator of the most contemporary study was selected to receive this survey. Site principal investigators were physicians in Pediatric Neurology or Pediatric Critical Care.

### Statistical analysis

Descriptive statistics were summarized as frequencies for categorical data and median and inter-quartile-range (IQR) for continuous data. Data from survey results were dichotomized based on the presence or absence of a clinical pathway for use of brain MRI as a prognostication tool following pediatric CA. Submitted surveys that did not include a response to this question were excluded from data analysis. Statistical analyses were performed using SAS^®^ software, Version 9.4 of the SAS System for Windows. Copyright © 2015, Cary, NC, USA.

## RESULTS

### Cohort characteristics

The survey was distributed to 143 individual institutions. Of those, 32% (46/143) completed the survey. Two surveys did not include a response to the survey question: “Does your center have a clinical pathway for the use of brain MRI as a prognostication tool following pediatric cardiac arrest?” and thus were excluded. The respondents included 27 institutions from the US (61.4%) and 17 international institutions (38.6%). Fifty-nine percent (26/44) of respondents were from freestanding children’s hospitals, 36% (16/44) were from hospitals annexed to an adult hospital, and 5% (2/44) responded “other” regarding hospital type. Participating sites had a median of 23 (IQR 12-36) pediatric ICU beds. A minority of sites (34% [15/44]) reported a clinical pathway to guide MRI use in CA. The number of CA cases at responding sites varied substantially (Table 1).

**Table 1.** Site characteristics.

| n(%) or median(25<sup>th</sup>, 75<sup>th</sup> percentiles) | All (n=44) | Clinical Pathway for MRI |  |
| --- | --- | --- | --- |
|  |  | No (n=29) | Yes (n=15) |
| Center's country | n=44 | n=29 | n=15 |
| United States | 27 (61.3) | 18 (62) | 9 (60) |
| Canada | 3 (6.8) | 3 (10.3) | 0 (0) |
| United Kingdom | 2 (4.5) | 1 (3.4) | 1 (6.67) |
| Spain | 3 (6.8) | 2 (6.9) | 1 (6.6) |
| Australia | 3 (6.8) | 2 (6.9) | 1 (6.6) |
| Other | 6 (13.6) | 3 (10.3) | 3 (20) |
| Type of hospital | n=44 | n=29 | n=15 |
| Freestanding children's hospital | 26 (59) | 15 (51.7) | 11 (73.3) |
| Children's hospital annexed to an adult hospital | 16 (36.3) | 13 (44.8) | 3 (20) |
| Other | 2 (4.55) | 1 (3.45) | 1 (6.67) |
| Number of PICU beds | n=43 | n=28 | n=15 |
|  | 23 (12 - 36) | 19 (13.5 - 32.5) | 36 (10 - 48) |
| Estimated no. pediatric in-hospital cardiac arrests per year | n=43 | n=29 | n=14 |
| < 10 | 10 (23.2) | 8 (27.5) | 2 (14.2) |
| 10-19 | 11 (25.5) | 9 (31.0) | 2 (14.2) |
| 20-29 | 12 (27.9) | 7 (24.1) | 5 (35.7) |
| 30-39 | 6 (13.9) | 2 (6.9) | 4 (28.0) |
| >= 40 | 3 (6.9) | 2 (6.9) | 1 (7.1) |
| Unknown | 1 (2.3) | 1 (3.4) | 0 (0.0) |
|  |  | No (n=29) | Yes (n=15) |
| Estimated no. pediatric out-of-hospital cardiac arrests per year | n=43 | n=28 | n=15 |
| < 10 | 11 (25.5) | 8 (28.5) | 3 (20.0) |
| 10-19 | 13 (30.2) | 9 (32.1) | 4 (26.6) |
| 20-29 | 13 (30.2) | 7 (25.0) | 6 (40.0) |
| 30-39 | 1 (2.3) | 1 (3.5) | 0 (0.0) |
| >= 40 | 4 (9.3) | 2 (7.1) | 2 (13.3) |
| Unknown | 1 (2.3) | 1 (3.5) | 0 (0.0) |

### Neuroimaging resources

The majority (75% [33/44]) of respondents reported that their institution employs a neuroradiologist to interpret brain MRIs (Table 2). Of centers with neuroradiology expertise, 55% (18/33) report these subspecialists are available to interpret MRIs 24/7. A higher percentage of institutions with a clinical pathway for post-CA MRI reported having 24/7 Neuro-radiology availability (57% vs. 34%). Eighty-one percent (35/43) of responding centers have a 3 Tesla MRI available, though 19% (8/43) are limited to 1.5 Tesla MRI.

**Table 2.**
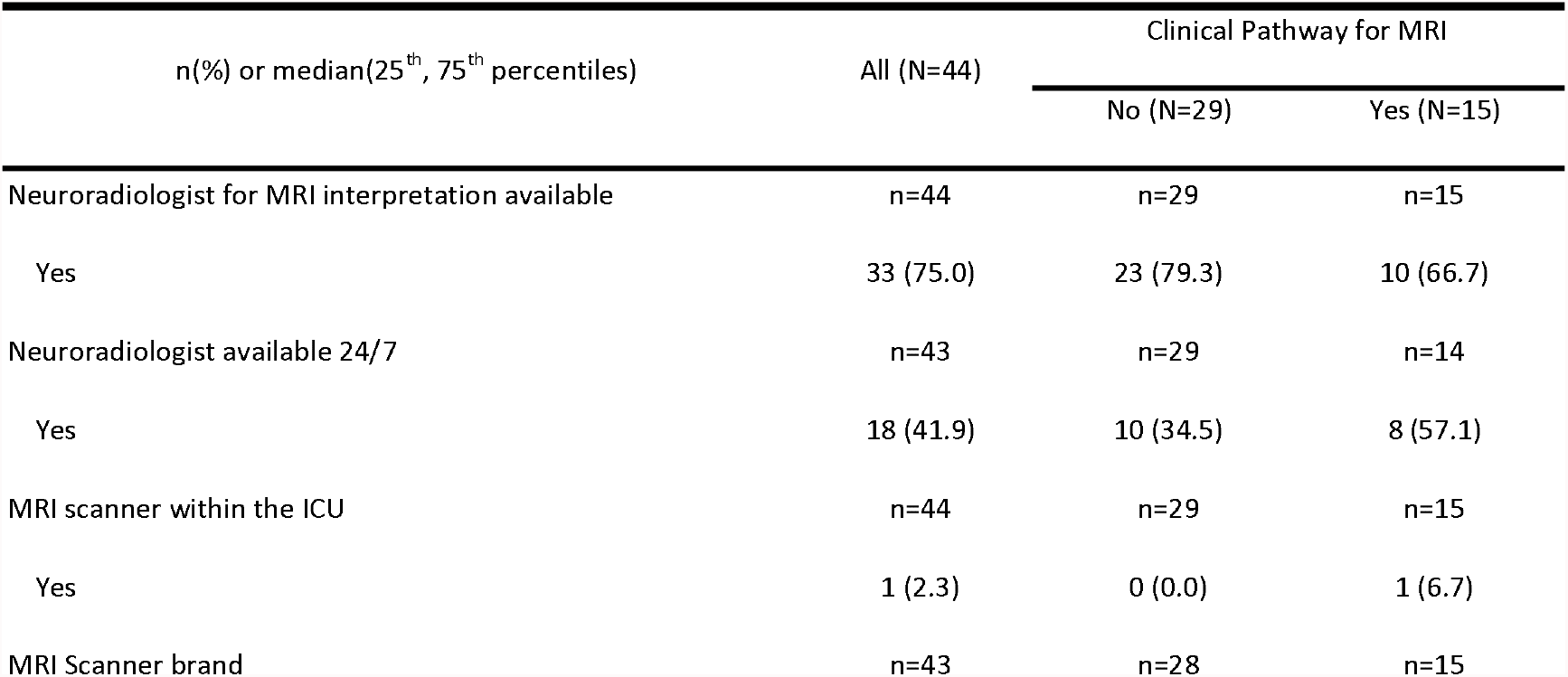
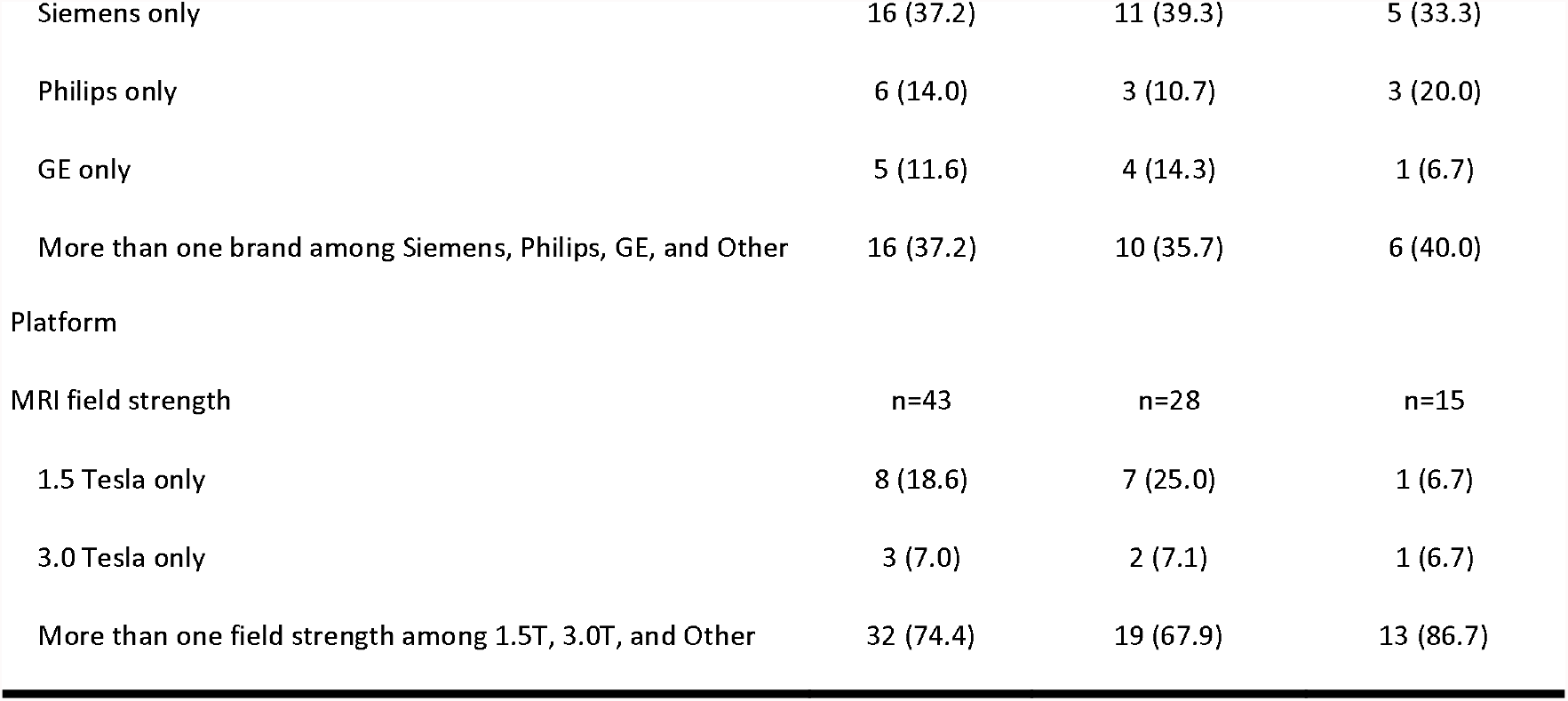
Neuroimaging resources.

| n(%) or median(25<sup>th</sup>, 75<sup>th</sup> percentiles) | All (N=44) | Clinical Pathway for MRI |  |
| --- | --- | --- | --- |
|  |  | No (N=29) | Yes (N=15) |
| Neuroradiologist for MRI interpretation available | n=44 | n=29 | n=15 |
| Yes | 33 (75.0) | 23 (79.3) | 10 (66.7) |
| Neuroradiologist available 24/7 | n=43 | n=29 | n=14 |
| Yes | 18 (41.9) | 10 (34.5) | 8 (57.1) |
| MRI scanner within the ICU | n=44 | n=29 | n=15 |
| Yes | 1 (2.3) | 0 (0.0) | 1 (6.7) |
| MRI Scanner brand | n=43 | n=28 | n=15 |
| Siemens only | 16 (37.2) | 11 (39.3) | 5 (33.3) |
| Philips only | 6 (14.0) | 3 (10.7) | 3 (20.0) |
| GE only | 5 (11.6) | 4 (14.3) | 1 (6.7) |
| More than one brand among Siemens, Philips, GE, and Other | 16 (37.2) | 10 (35.7) | 6 (40.0) |
| Platform |  |  |  |
| MRI field strength | n=43 | n=28 | n=15 |
| 1.5 Tesla only | 8 (18.6) | 7 (25.0) | 1 (6.7) |
| 3.0 Tesla only | 3 (7.0) | 2 (7.1) | 1 (6.7) |
| More than one field strength among 1.5T, 3.0T, and Other | 32 (74.4) | 19 (67.9) | 13 (86.7) |

### Practices regarding MRI use in cardiac arrest

Of the participating institutions, 45% (20/44) have a dedicated Pediatric Neurocritical Care team/service (Table 3). Institutions with a pediatric neurocritical care team/service were almost twice as likely to have a clinical pathway for MRI after CA (66.7% vs. 34.5%). Fifty percent of institutions (22/44) obtain an MRI in nearly all pediatric CA patients and 31.8% (14/44) of institutions reported performing MRI in patients who are not back to their baseline. Centers with a clinical pathway for MRI use after CA were less likely to obtain an MRI on a “case by case” basis (6.7% [1/15]) compared to centers without a clinical pathway (24.1% [7/29]).

**Table 3.**
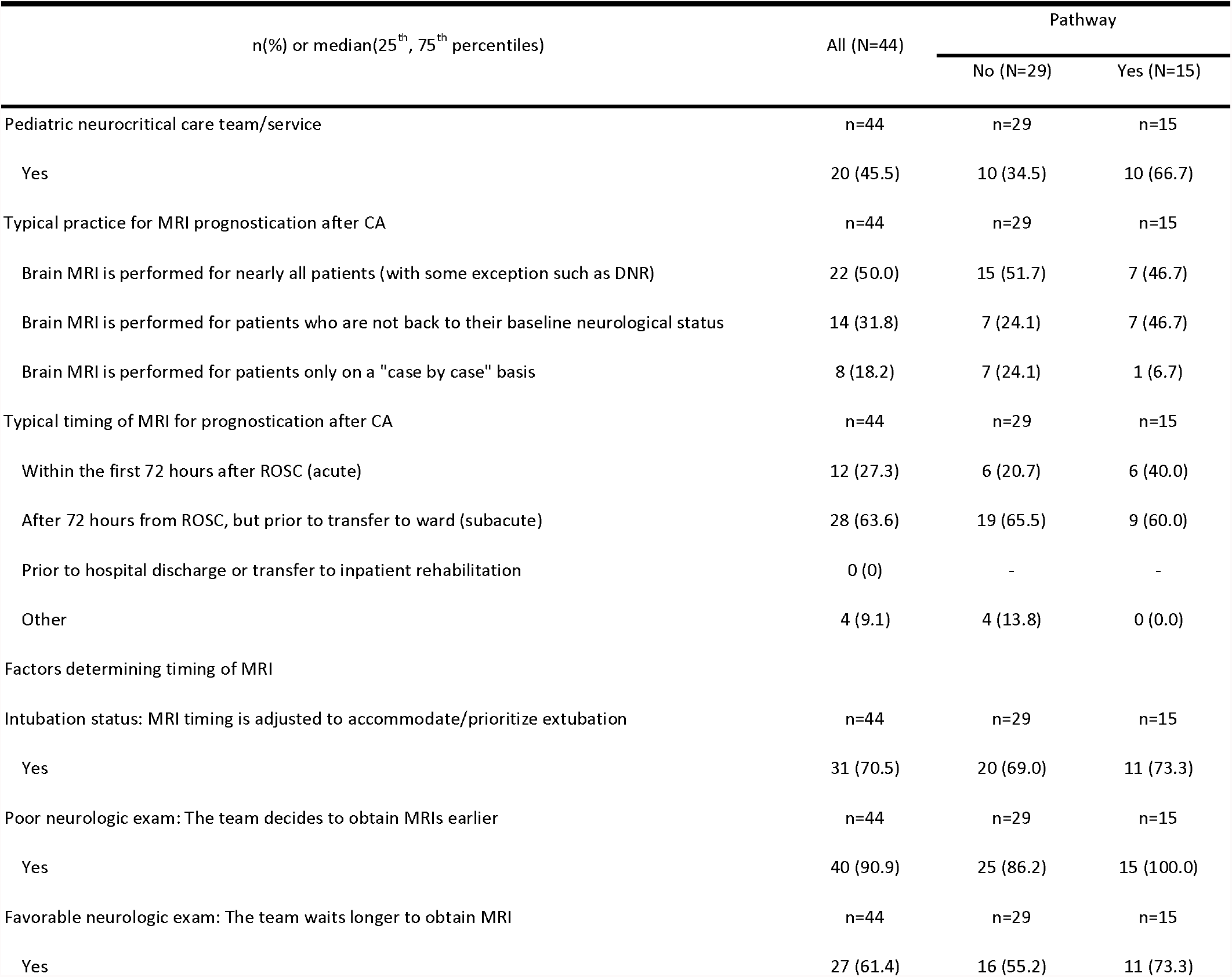
Clinical practices regarding the use of MRI in pediatric CA.

| n(%) or median(25<sup>th</sup>, 75<sup>th</sup> percentiles) | All (N=44) | Pathway |  |
| --- | --- | --- | --- |
|  |  | No (N=29) | Yes (N=15) |
| Pediatric neurocritical care team/service | n=44 | n=29 | n=15 |
| Yes | 20 (45.5) | 10 (34.5) | 10 (66.7) |
| Typical practice for MRI prognostication after CA | n=44 | n=29 | n=15 |
| Brain MRI is performed for nearly all patients (with some exception such as DNR) | 22 (50.0) | 15 (51.7) | 7 (46.7) |
| Brain MRI is performed for patients who are not back to their baseline neurological status | 14 (31.8) | 7 (24.1) | 7 (46.7) |
| Brain MRI is performed for patients only on a "case by case" basis | 8 (18.2) | 7 (24.1) | 1 (6.7) |
| Typical timing of MRI for prognostication after CA | n=44 | n=29 | n=15 |
| Within the first 72 hours after ROSC (acute) | 12 (27.3) | 6 (20.7) | 6 (40.0) |
| After 72 hours from ROSC, but prior to transfer to ward (subacute) | 28 (63.6) | 19 (65.5) | 9 (60.0) |
| Prior to hospital discharge or transfer to inpatient rehabilitation | 0 (0) | - | - |
| Other | 4 (9.1) | 4 (13.8) | 0 (0.0) |
| Factors determining timing of MRI |  |  |  |
| Intubation status: MRI timing is adjusted to accommodate/prioritize extubation | n=44 | n=29 | n=15 |
| Yes | 31 (70.5) | 20 (69.0) | 11 (73.3) |
| Poor neurologic exam: The team decides to obtain MRIs earlier | n=44 | n=29 | n=15 |
| Yes | 40 (90.9) | 25 (86.2) | 15 (100.0) |
| Favorable neurologic exam: The team waits longer to obtain MRI | n=44 | n=29 | n=15 |
| Yes | 27 (61.4) | 16 (55.2) | 11 (73.3) |

The majority of institutions (63.6% [28/44]) perform an MRI after 72 hours of return of spontaneous circulation (ROSC) but before transfer to the inpatient ward, 27.3% of institutions (12/44) obtain an MRI within 72 hours of ROSC and (9% (4/44) report the timing of MRI for prognostication after CA as “other.” All centers with a clinical pathway reported obtaining an MRI either within the first 72 hours after ROSC (40% [6/15]) or greater than 72 hours after ROSC and before transfer to the inpatient ward (60% [9/15]). None of the centers indicating the timing of MRI as “other” reported having a clinical pathway. When asked to indicate what factors are used to determine timing of brain MRI for prognostication, 90% of centers (40/44) reported poor neurologic exam as the main factor determining the timing of the MRI. Other clinical factors determining timing of imaging for prognostication included intubation status (70% [31/44]) and favorable neurologic exam (61% [27/44]). All centers with a clinical pathway for MRI after CA indicated at least one clinical factor is used to determine timing of imaging for prognostication. Only one center reported using no clinical factors to determine timing of MRI.

### Types of MRI sequences used in children with cardiac arrest

Sites were asked to indicate which sequences were included in the imaging protocol used to evaluate children with CA. The percentage of centers using each specific MRI sequence, stratified by centers with and without a pathway, is illustrated in Figure 1. Over 97% (43/44) of institutions use T1, T2, and fluid-attenuated inversion recovery (FLAIR). All centers reported obtaining diffusion-weighted imaging (DWI). Sequences sensitive to hemorrhage such as gradient-echo (GRE) and susceptibility weighted imaging (SWI) are used in 43% and 67% of centers, respectively. Less than half of the institutions (40% and 31% of institutions with and without a pathway, respectively) report use of spectroscopy (MRS). Other techniques, such as functional MRI, MR venogram, and post-contrast T1 are used by less than 30% of institutions, regardless of the presence or absence of a pathway.

**Figure 1.**
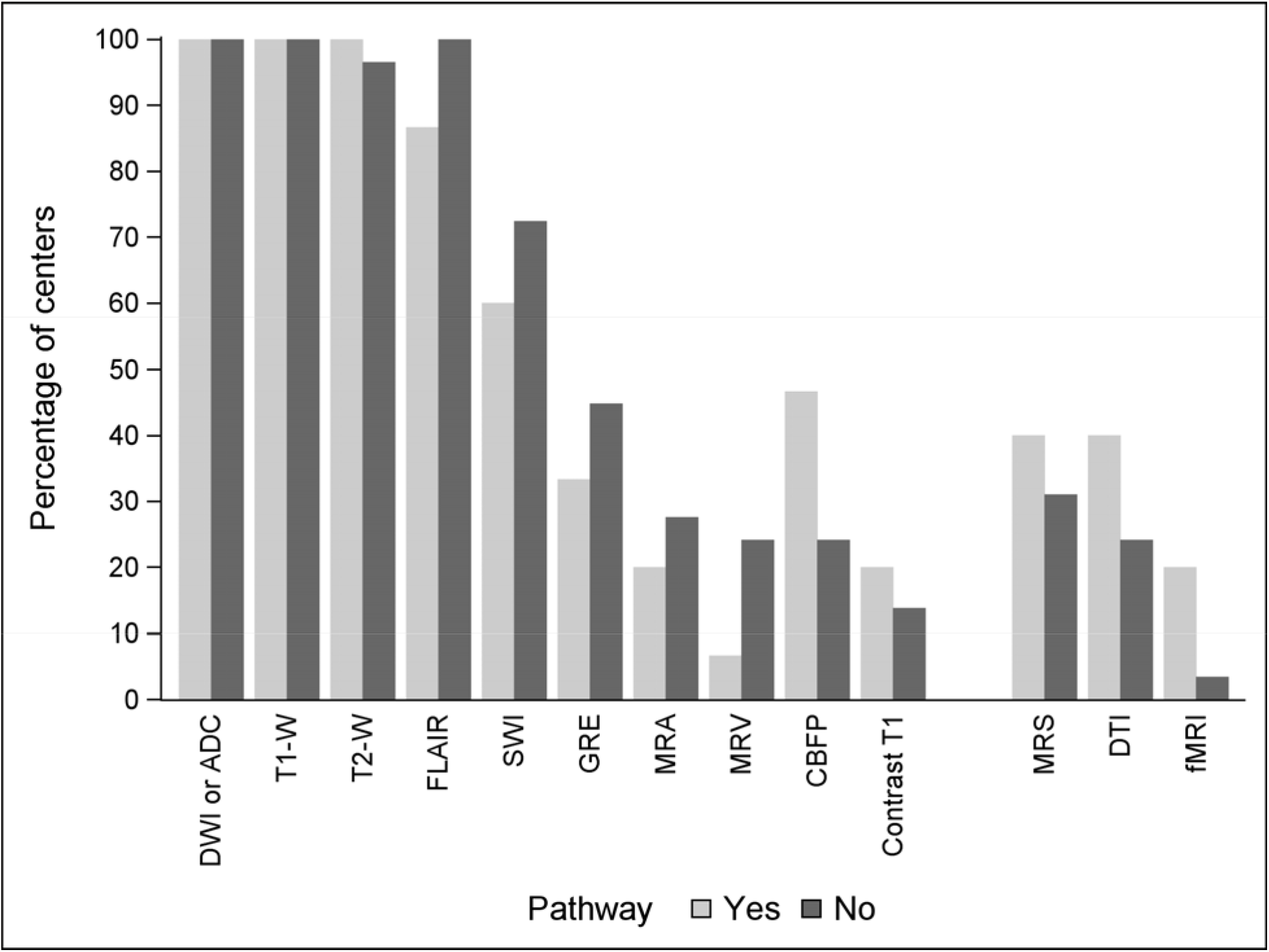
Type of MRI sequences used in pediatric cardiac arrest Figure 1 Definitions: DWI-Diffusion weighted imaging; ADC-Apparent diffusion coefficient; FLAIR-Fluid-attenuated inversion recovery; GRE-Gradient Echo; SWI-Susceptibility weighted imaging; CBFP-Cerebral blood flow/perfusion; MRS-Magnetic resonance spectroscopy; DTI-Diffusion tensor imaging; MRA-Magnetic resonance angiogram; MRV-Magnetic resonance venogram; fMRI-Resting state functional MRI.

## DISCUSSION

Our study describes current practices in use of brain MRI as a neuroprognostic tool in children with CA from 44 national and international pediatric tertiary care centers. The results suggest that clinicians often perform MRI as part of their routine care in children with CA, with half of responding institutions indicating that they obtain MRI in nearly all patients post-CA, and an additional 30% indicating that they obtain an MRI in those who are not back to their neurological baseline. Nearly all institutions routinely incorporate conventional sequences (i.e., T1, T2, DWI and FLAIR). Reported use of sequences sensitive to hemorrhage are 43% (GRE) and 67% (SWI). These rates appear low, but it is likely that centers choose to utilize a single sequence to evaluate for hemorrhage, resulting in little overlap in these values, thus indicating that this practice is also widely used at surveyed centers. However, there is still significant heterogeneity and less use of advanced MRI sequences post CA. We found that only a minority of the protocols incorporate advanced imaging sequences such as MRS. There are several potential reasons or explanations that may contribute to these observations. First, although promising, data regarding the clinical value of sequences such as MRS is limited to single-center, retrospective studies. Second, the interpretation of these images requires particular expertise. In our study, 25% of centers do not have a member of the radiology team with specific training in neuroradiology.

In our study, only 34% of centers indicated that they have an established clinical pathway for the use of MRI in pediatric CA. Institutions with CA pathways informing MRI utilization reported less variation in patient selection or timing of MRIs. The lack of pathway-driven practices is likely due to a number of factors including: sparse and low quality evidence in pediatrics and practice guidelines developed by data extrapolated from adult patients or by expert opinion.^31^ Additionally, an increasing number of institutions have incorporated pediatric neurocritical care teams to build institutional expertise and provide standardized, evidence-based care for children with acute brain injury.^32, 33^ While less than half of responding institutions have a Pediatric Neurocritical Care team available, the presence of a Pediatric Neurocritical Care team doubled the likelihood of pathway driven care.

Taken together, these observations further highlight the need for evidence to guide clinical practice in the area of pediatric CA. Recent guidelines published by the American Heart Association (AHA) cite insufficient evidence to support the use of MRI as a standard of care in pediatric CA and state that “MRI during the first seven days post-arrest may be valuable” in assisting with prognostication.^12^ Beyond this, no recommendations are made regarding indications, optimal timing, or acquisition protocol. The guidelines cite retrospective study design, small sample size, selection bias, lack of blinding in imaging interpretation, variable outcome measures, and follow-up times as limitations to the current evidence.

Conventional MRI protocols designed for clinical use include sequences such as T1 and T2-weighted images, FLAIR and diffusion-weighted imaging.^25^ Prior studies have investigated the utility of these sequences in the assessment of children with CA. In a single-center study of 28 children who received a brain MRI within the first 14 days post-ROSC, T2 signal intensity abnormalities in the basal ganglia and diffusion restriction in the cerebral lobes were associated with unfavorable outcomes.^15^ A separate study evaluated the correlation between imaging abnormalities and unfavorable outcomes in 23 children with CA who underwent MRI within 14 days after ROSC.^10^ The injured areas most commonly identified by T2-weighted imaging were the lentiform nucleus (68%), the caudate (55%), and the thalamus (50%). T2 abnormalities in the lentiform nuclei were associated with unfavorable outcomes at hospital discharge. Diffusion restriction was most commonly seen in the lentiform nucleus (41%), thalamus (32%), frontal (30%), and parietal (30%) lobes. More recently, a single center study of 77 pediatric patients after OHCA who had an MRI at a median of day 4 post-CA found that the extent of diffusion restriction and T2/Flair abnormalities were associated with unfavorable outcomes with good discrimination (AUROC of 0.96 (95% CI 0.91, 0.99) for diffusion and 0.92 (0.85, 0.97) for T2/FLAIR injury).^17^ In addition to T2-weighted and DWI, more advanced imaging modalities such as MRS may have prognostic value in this population. Increased lactate concentrations in the parietooccipital gray matter and decreased NAA in the parietal white matter using MRS have been observed in children with CA and unfavorable outcomes.^10^ Decreased NAA and increased lactate were also observed in a study of children with acute brain injury (including CA) and unfavorable outcomes.^20^

Given the evolution of the appearance of brain lesions on MRI, both the composition of the MRI protocol and the timing of the initial MRI after presentation are critical to capture the extent of brain injury accurately. MRI sequences sensitive to ischemia and cytotoxic edema such as DWI/apparent diffusion coefficient (ADC) often identify lesions in children with CA. Unlike abnormalities on T1 and T2 MRI sequences, diffusion abnormalities can be seen within minutes in acute ischemia (i.e. arterial ischemic stroke). However, diffusion abnormalities can also be delayed in CA possibly due to the variability of hemodynamic compromise over the course of the resuscitation and post resuscitation period.^26-29^ Thus, to ensure appropriate evaluation of the presence or absence of hypoxic ischemic brain injury after CA, the timing of MRI is very important. The AHA guidelines recommend considering use of MRI “within the first seven days” after injury.”^12^ In our survey, most centers stated that they perform MRIs after 72 hours post-ROSC. Ninety percent stated that poor neurologic exam is a main determinant for obtaining an MRI early and 61% stated that a favorable neurologic exam would lead to clinical teams waiting longer to obtain an MRI.

Our survey has a number of limitations that should be considered when interpreting these results. First, our survey response rate was relatively low at 30%. Second, a limitation common to survey studies with a single respondent per center is that the responses are based on the individual’s understanding of current institutional practices and may represent her/his opinion and practice over fact. Third, the surveyed institutions represent tertiary care facilities with an interest in clinical research in pediatric cardiac arrest which may not be generalizable to all pediatric intensive care settings. Imaging resources and practices may differ from institutions not participating in large research consortia, limiting the external validity of our findings. Of note, the cohort was not limited to large centers and included institutions with a broad spectrum of size and number of CA cases per year. Lastly, the correlation between the indications for MRI, timing, and acquisition protocols and patient outcomes was beyond the scope of this survey. Prospective, adequately powered, multi-center studies with standardized, patient-oriented outcomes are needed to bridge this knowledge gap.

Our survey reveals prevalent use of MRI for neuroprognostication in pediatric CA. Responses suggest a wide variability in the timing, MRI sequences, and neuroradiology resources, as well as a dearth of centers with clinical pathways to guide MRI utilization after pediatric CA. Studies to ultimately determine the optimal use of MRI in pediatric CA will need to account for and implement strategies reduce this variation.

## Supporting information

Survey

## Data Availability

All data produced in the present study are available upon reasonable request to the authors

## Acknowledgements

We acknowledge members of the **POCCA Steering Committee**: Patrick Kochanek, MD, MCCM; Robert Clark, MD; Hulya Bayir, MD; Ashok Panigrahy, MD; Rachel Berger, MD, MPH; Sue Beers, PhD; Tony Fabio, PhD, MPH; **POCCA Investigators**: Karen Walson, MD (Children’s Healthcare of Atlanta); Alexis Topjian, MD, MSCE (Children’s Hospital of Philadelphia); Christopher JL Newth, MD, FRCPC (Children’s Hospital of Los Angeles); Elizabeth Hunt, MD, MPH, PhD, Jordan Duval-Arnould, DrPH, MPH (Johns Hopkins Children’s Center); Binod Balakrishnan, MD, Michael T. Meyer, MD, MS, FCCM (Children’s Hospital of Wisconsin); Melissa G. Chung, MD (Nationwide Children’s Hospital); Anthony Willyerd, MD (Phoenix Children’s Hospital); Lincoln Smith, MD, Jesse Wenger, MD (Seattle Children’s); Stuart Friess, MD, Jose Pineda, MD, MS (St. Louis Children’s); Ashley Siems, MD, Jason Patregnani, MD, John Diddle, MD (Children’s National Hospital); Aline Maddux, MD, MSCS and Craig Press, MD, PhD (Children’s Hospital of Colorado); Lesley Doughty, MD (Cincinnati Children’s Hospital Medical Center); Juan Piantino, MD (Doernbecher Children’s Hospital); **and the POCCA Coordinators**: David Maloney, BS, Pamela Rubin, RN (UPMC Children’s Hospital of Pittsburgh); Beena Desai, BS, CCRC, Maureen G. Richardson, BSN, RN, CPN, Cynthia Bates, CCRP (Children’s Healthcare of Atlanta); Darshana Parikh, Janice Prodell, Maddie Winters, Katherine Smith, MPH, BSN, RN, CPN (Children’s Hospital of Philadelphia); Jeni Kwok, JD, Adriana Cabrales, BA (Children’s Hospital of Los Angeles); Ronke Adewale, Pam Melvin(Johns Hopkins Children’s Center); Sadaf Shad, Katherine Siegel, Katherine Murkowski, Mary Kasch (Children’s Hospital of Wisconsin); Josey Hensley RN, BSN, Lisa Steele, RN, BSN (Nationwide Children’s Hospital); Danielle Brown, Brian Burrows, Lauren Hlivka (Phoenix Children’s Hospital); Deana Rich (Seattle Children’s Hospital); Amila Tutundzic, Tina Day, Lori Barganier (St. Louis Children’s Hospital); Ashley Wolfe, Mackenzie Little, Elyse Tomanio, Neha Patel, Diane Hession (Children’s National Hospital); Yamila Sierra MPH, CCRP (Children’s Hospital of Colorado); and Rhonda Jones, Laura Benken (Cincinnati Children’s).

We also thank Jonathan Elmer, MD, MS; Subramanian Subramanian, MD; Srikala Narayanan, MD; Julia Wallace; Tami Robinson; Andrew Frank; Stefan Bluml, PhD; Jessica Wisnowski, PhD; Keri Feldman; Avinash Vemulapalli; Linda Ryan; Scott Szypulski, MBA; Christopher Keys, and all of the children and families who generously participated in POCCA. We are grateful to the all ICU staff, nurses, and physicians for their efforts in study recruitment and provision of excellent clinical care to families.

