## Supplementary material for "Use of magnetic resonance imaging in neuroprognostication after pediatric cardiac arrest: Survey of current practices": Survey

**Objective**: The main goal of this study is to describe current practices regarding the use of brain MRI for prognostication in children after cardiac arrest.

**Patient population:** Hospitalized children receiving post-cardiac arrest care after an in- or out-of-hospital cardiac arrest.

**Instructions**: Answers should reflect your center's clinical care pathway, or in the case where there is no pathway, the most common practices employed. The survey will take approximately 5-10 minutes to complete. Consultation with a radiology or neurology colleague who is knowledgeable about MRI practices in your center is encouraged (or permissible) if needed.

### SECTION I: SITE CHARACTERISTICS

1. Full name of your center: ______________________________

2. Your center's country:
   1 ☐ United States　2 ☐ Canada　3 ☐ United Kingdom
   4 ☐ Spain　5 ☐ Netherlands　6 ☐ New Zealand
   7 ☐ India　8 ☐ Australia　9 ☐ South Africa
   10 ☐ Other: ______________________________

3. Indicate the type of hospital and record the number of beds for children:

   3.1 Type of hospital:
   1 ☐ Freestanding children's hospital
   2 ☐ Children's hospital annexed to an adult hospital
   3 ☐ Other:

   3.2 Number of pediatric hospital beds in your center: ______

4. Indicate each type of ICU in your center that care for children (not NICU) with number of beds *(check all that apply)*:

   ☐ General PICU: Number of beds: ______
   ☐ Pediatric Cardiac ICU: Number of beds: ______
   ☐ Other ICU: ______________________________: Number of beds: ______

5. Does your center have a pediatric neurocritical care team/service: ☐ Yes ☐ No

   5.1 If yes, does your center employ either of the following for post-cardiac arrest resuscitation patient care:
   1 ☐ Pediatric neurocritical care team acting as primary providers
   2 ☐ Pediatric neurocritical care consultation service
   3 ☐ Both

6. Indicate the estimated number of pediatric cardiac arrest patients cared for at your center per year in your PICU, CICU, and other ICU combined (not NICU):

   6.1 In-hospital cardiac arrest: 1 ☐ < 10　2 ☐ 10-19　3 ☐ 20-29　4 ☐ 30-39　5 ☐ ≥ 40
   6.2 Out of hospital cardiac arrest: 1 ☐ < 10　2 ☐ 10-19　3 ☐ 20-29　4 ☐ 30-39　5 ☐ ≥ 40

**Use of Magnetic Resonance Imaging in Pediatric Cardiac Arrest Survey**

**SECTION I: SITE CHARACTERISTICS *(Continued)***

7. Is there a dedicated neuroradiologist who provides brain MRI interpretation for children with cardiac arrest at your center?
   - ☐ Yes
     If yes,
     7.1 Is the neuroradiologist available to provide MRI reads 24 hours a day/7 days a week at your center? ☐ Yes ☐ No
   - ☐ No
     If no,
     7.2 What is the training background of the person responsible for providing the MRI reads at your center: [ ]

8. Provide the number of MRI scanners available in your center by platform and magnet field strength *(check all that apply)*:
   - ☐ Siemens:
     9.1 Field strength *(check all that apply)*:
     - ☐ 1.5 T: Number of MRI scanners: ______
     - ☐ 3.0 T: Number of MRI scanners: ______
     - ☐ Other: ______________________ Number of MRI scanners: ______
   - ☐ General Electric:
     9.2 Field strength *(check all that apply)*:
     - ☐ 1.5 T: Number of MRI scanners: ______
     - ☐ 3.0 T: Number of MRI scanners: ______
     - ☐ Other: ______________________ Number of MRI scanners: ______
   - ☐ Philips
     9.3 Field strength *(check all that apply)*:
     - ☐ 1.5 T: Number of MRI scanners: ______
     - ☐ 3.0 T: Number of MRI scanners: ______
     - ☐ Other: ______________________ Number of MRI scanners: ______
   - ☐ Other Platform 1:
     9.4 Field strength *(check all that apply)*:
     - ☐ 1.5 T: Number of MRI scanners: ______
     - ☐ 3.0 T: Number of MRI scanners: ______
     - ☐ Other: ______________________ Number of MRI scanners: ______
   - ☐ Other Platform 2:
     9.5 Field strength *(check all that apply)*:
     - ☐ 1.5 T: Number of MRI scanners: ______
     - ☐ 3.0 T: Number of MRI scanners: ______
     - ☐ Other: ______________________ Number of MRI scanners: ______

**Use of Magnetic Resonance Imaging in Pediatric Cardiac Arrest Survey**

**SECTION I: SITE CHARACTERISTICS *(Continued)***

9. Which type of MRI scanner is most commonly used in children with cardiac arrest?
   1 ☐ Siemens 1.5T    2 ☐ Siemens 3T    3 ☐ General Electric 1.5T    4 ☐ General Electric 3.0T
   5 ☐ Philips 1.5T    6 ☐ Philips 3.0T    7 ☐ Other

10. Where are your MRI scanners located *(check all that apply)*?
   ☐ Within the ICU: Number in this location: ______
   ☐ Outside of the ICU – same floor: Number in this location: ______
   ☐ Outside of the ICU – different floor: Number in this location: ______

**SECTION II: APPROACH TO MRI**

1. Does your center have a clinical pathway for the use of brain MRI as a prognostication tool following pediatric cardiac arrest?
   ☐ Yes    ☐ No

2. Of the following sentences, which one most accurately describes your center's typical practice regarding the use of MRI for prognostication after pediatric cardiac arrest?
   1 ☐ Brain MRI is performed for nearly all patients (with some exception such as DNR).
   2 ☐ Brain MRI is performed for patients who are not back to their baseline neurological status.
   3 ☐ Brain MRI is performed for patients only on a "case by case" basis.

3. What is the typical timing of brain MRI for prognostication purposes following pediatric cardiac arrest at your center?
   1 ☐ Within the first 72 hours after ROSC (acute)
   2 ☐ After 72 hours from ROSC, but prior to transfer to ward (subacute)
   3 ☐ When the child is ready for hospital discharge/inpatient rehabilitation (later)
   4 ☐ Other: ________________

**Use of Magnetic Resonance Imaging in Pediatric Cardiac Arrest Survey**

**SECTION II: APPROACH TO MRI *(Continued)***

4. What MRI sequences are included in your center's clinical care pathway or are typically used if you do not have a pathway for prognostication post-arrest? (*check all that apply*)

☐ T1 weighted imaging
☐ T2 weighted imaging
☐ Fluid-attenuated Inversion recovery (FLAIR)
☐ Diffusion weighted imaging (DWI)/Apparent diffusion coefficient (ADC)
☐ MR Spectroscopy
☐ Cerebral blood flow/perfusion → Approach:
  ☐ Arterial Spin Labeling (ASL) perfusion
  ☐ Dynamic Susceptibility Contrast (DSC) perfusion
☐ Diffusion tensor imaging (DTI)
☐ Resting state functional MRI (fMRI)
☐ Susceptibility Weighted Imaging (SWI)
☐ Gradient echo (GRE)
☐ Contrast T1
☐ MR angiogram (MRA)
☐ MR venogram (MRV)
☐ Other: ______________________________

5. When determining the timing of MRI for prognostication after cardiac arrest, what factors could inform your center's practice *(check all that apply)*:

☐ Intubation status (I.e. MRI timing is adjusted to accommodate/prioritize extubation)
☐ Poor neurologic exam (i.e. the team decides to obtain MRIs earlier)
☐ Favorable neurologic exam (i.e. the team waits longer to obtain MRI)
☐ None (MRI is obtained per protocol no matter what the patient's clinic status is)

6. Besides prognostication what other purposes is brain MRI used for in pediatric cardiac arrest patients at your center? *(check all that apply)*

☐ Diagnosis of cardiac arrest etiology
☐ Explanation of persistent altered mental status
☐ Evaluation of etiology of seizures
☐ Other: ______________________________
